# Paracingulate sulcus length is shorter in voice-hearers regardless of need for care

**DOI:** 10.1101/2020.01.13.19015891

**Authors:** Albert R. Powers, Laura I. van Dyck, Jane R. Garrison, Philip R. Corlett

## Abstract

While hallucinations are often considered to be the sine qua non of serious mental illness, they are commonly experienced by those without a need for care. Such non-clinical, hallucinating individuals offer a window into the etiology of hallucinations. Garrison et al. (2015) recently assessed structural variability in paracingulate sulcus (PCS) of medial prefrontal cortex. They found a reduction of 10mm in left hemisphere PCS length increased the likelihood of hallucinations by 19.9%. However, these findings were in clinical hallucinators, and reduced leftward asymmetry in PCS has been repeatedly demonstrated in schizophrenia and associated with reduced reality monitoring. Similar analyses have been conducted in non-clinical voice hearers and in those particpants, no reduction in PCS length was reported. However, there is considerable variability in the voice experiences of non-clinical voice hearers. Put simply, some may be more clinical than others. We recently identified a relatively homogenous group of non-clinical voice hearers who identify as clairaudient psychics. Their voice experiences are extremely redolent of those reported by a matched patient voice hearing group and their brain and behavioral responses to a perceptual inference task were, at least at the level of prior beliefs, similarly perturbed to patients who hear voices. These observations are consistent with a continuum model of voice hearing, along which clairaudient voice hearers are nearer to patients with schizophrenia in some respects. The PCS data reported by Garrison et al would militate against such a continuum. Before rejecting the continuum model, we sought to determine if the differences in PCS length also applied to the clairaudient non-clinical voice hearers. If PCS length is to be developed as a biomarker for clinical hallucination risk, then it is important to demonstrate that it is not similarly reduced in non-clinical voice hearers. We find that it is reduced, both in clinical and non-clinical voice-hearers.

## Introduction

While hallucinations are often considered to be the *sine qua non* of serious mental illness, they are commonly experienced by those without a need for care^1^. Such non-clinical, hallucinating individuals offer a window into the etiology of hallucinations.

Garrison et al. (2015) recently assessed structural variability in paracingulate sulcus (PCS) of medial prefrontal cortex. They found a reduction of 10mm in left hemisphere PCS length increased the likelihood of hallucinations by 19.9%^2^. However, these findings were in clinical hallucinators, and reduced leftward asymmetry in PCS has been repeatedly demonstrated in schizophrenia and associated with reduced reality monitoring^3^. Similar analyses have been conducted in non-clinical voice hearers and in those particpants, no reduction in PCS length was reported^4^. However, there is considerable variability in the voice experiences of non-clinical voice hearers. Put simply, some may be more clinical than others. We recently identified a relatively homogenous group of non-clinical voice hearers who identify as clairaudient psychics√. Their voice experiences are extremely redolent of those reported by a matched patient voice hearing group^5^ and their brain and behavioral responses to a perceptual inference task were, at least at the level of prior beliefs, similarly perturbed to patients who hear voices^6^. These observations are consistent with a continuum model of voice hearing, along which clairaudient voice hearers are nearer to patients with schizophrenia in some respects. The PCS data reported by Garrison et al would militate against such a continuum^4^. Before rejecting the continuum model, we sought to determine if the differences in PCS length also applied to the clairaudient non-clinical voice hearers.

If PCS length is to be developed as a biomarker for clinical hallucination risk, then it is important to demonstrate that it is not similarly reduced in non-clinical voice hearers.

## Methods

Recruitment, clinical assessment, and image acquisition are detailed in Powers et al. (2017). The project was approved by the local Institutional Review Board and conducted in accordance with the Declaration of Helsinki. T1-weighted structural MRI scans were obtained using 3D MPRAGE sequence for four groups of participants: 1) voice-hearers with a psychotic disorder (P+H+; n = 15); 2) voice-hearers without a psychotic disorder (P-H+; n = 15); 3) non-voice-hearers with a psychotic disorder (P+H-; n = 14); and 4) non-voice-hearers without a psychotic disorder (P-H-; n = 15). All analyses were carried out blind to participant group.

Scans were imported into and calibrated using Mango brain visualization software (version 3.6). Using sagittal slices, 4mm to the left or right of the medial line, the PCS was identified and measured following prior work^2^. The full protocol for measuring PCS length is available here: https://www.repository.cam.ac.uk/handle/1810/264520. Briefly, individual scans were imported as NIFTI folders into Mango brain visualization software (version 3.6; http://ric.uthscsa.edu/mango) and inspected for integrity. In an axial view, the locations of the anterior and posterior commissures (AC and PC) were marked, and the scan rotated to line up the AC and PC in a horizontal plane. The origin was reset to the location of the AC. On a sagittal slice, 4.375mm to the left or right of the medial line, the cingulate sulcus (CS) was identified as the first major sulcus running in an anterior– posterior direction, dorsal to the corpus callosum and typically visible for five sagittal slices or more. The PCS was then identified if present as a salient sulcus, running parallel, horizontal and dorsal to the CS, and visible for three or more sagittal slices measured from the medial plane (x = 0). The sulcus was measured using the ‘trace line’ function in Mango from its start in the first quadrant prescribed by y > 0 and the horizontal line linking the AC and PC (z > 0), starting at the point at which the sulcus ran in a posterior direction. The PCS was measured to its end point, which could fall outside of the first quadrant All analyses were conducted in MATLAB 2014b.

## Results

With two-way ANOVA, there was a main effect of hallucination status—but not psychosis—on right PCS length, with no interaction of hallucinations and psychosis. This effect was not seen in the left PCS. Participants with hallucinations exhibited reduced right PCS length compared to participants without hallucinations (mean reduction = 8.8 mm, P<0.05). The effect of hallucinations on right PCS length was sufficiently robust to persist with the addition to the model of the potentially confounding covariates: age and IQ (F =4.55, P< 0.05).

## Discussion

While past studies have demonstrated decreased PCS length in individuals with psychosis and hallucinations compared to their non-hallucinating counterparts^2^, ours is the first demonstration that such an effect extends to non-treatment-seeking voice-hearers. Right PCS in our sample was significantly shorter in voice-hearers, regardless of whether or not they had a diagnosable psychotic illness. In fact, mean right PCS length was numerically shorter for non-clinical voice-hearers than for any other group, although this difference did not reach statistical significance. Our findings support the hypothesis that differences in PCS length are specifically related to the propensity to hear voices and not to illness, disability, or dysfunction^2^. However, our non-clinical voice hearers’ experiences were very redolent of those reported by patients with schizophrenia^5^. It is possible that not all non-clinical voice hearers have experiences— and, we would argue—brains - that match patients so closely.

We recently demonstrated a hallucination-specific effect in an adjacent prefrontal region during conditioned hallucinations^6^. We showed that this region’s activity encodes low-level perceptual beliefs about the sensory environment that may drive hallucinations. It may be that the morphological differences highlighted here play a role in mediating perceptual belief formation or its interactions with incoming sensory evidence during hallucinations. Recently-identified methods for functional MRI at the resolution of cortical layers (i.e., laminar fMRI^7^) may aid in fully characterizing these links between PCS morphology and the computational underpinnings of hallucinatory phenomena.

**Figure 1.**
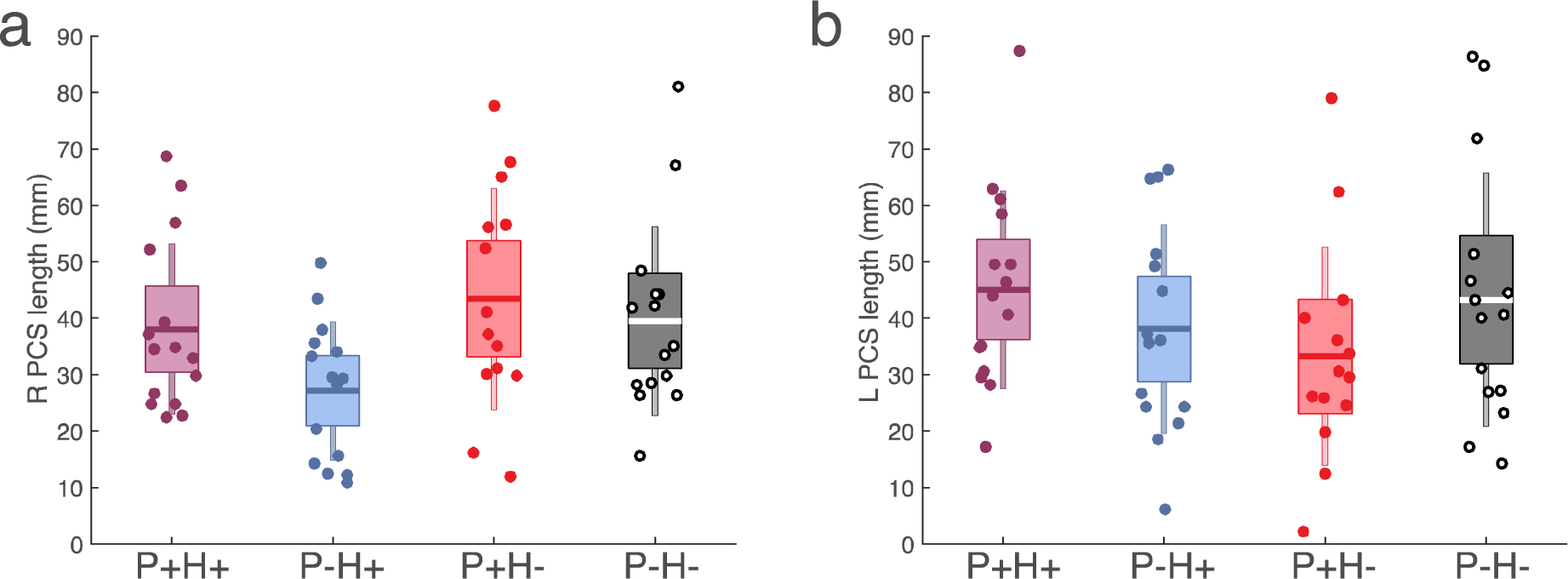
Paracingulate sulcus (PCS) length. Right PCS is significantly shorter in participants with hallucinations than in participants without (**a**), but this effect is not seen in left PCS (**b**).

## Data Availability

Imaging data are stored at NeuroVault (http://neurovault.org/collections/OCFEJCQE/).

